# Impact of Donor History on the Risk of Transfusion-Related Infections

**DOI:** 10.1101/2025.01.03.25319941

**Authors:** Collince Odiwuor Ogolla, Benard Guya, Apollo O. Maima

## Abstract

**Background:** Transfusion-related infections are a severe threat to the safety of transfusing blood products, internationally. Advances in screening procedures have not, nevertheless, rendered blood transfusion a risk-free procedure for transmitting infectious disease. Donor history, including previous donation history, medical conditions, and high-risk behavior, may influence transfusion-related infections.

**Objective:** The objective of this study was to determine donors’ histories that could influence the possibility of transfusion-related infections.

**Methods:** The cross-sectional study was done in January 2022 to December 2023 at KTRH. Out of 108 blood transfusions, data for 108 patients were retrieved from patient medical records and donor screening forms. Variables such as HIV and Hepatitis among donor medical conditions, previous donation history, and high-risk behaviors such as intravenous drug use were analyzed for their link with TRIs. The diagnosis of TRIs was established based on the results of a clinical examination and laboratory tests. Using descriptive statistics as well as Chi-square tests and logistic regression, data were analyzed.

**Results:** Of the total transfusion sample (n=108), 15 (13.9%) recipients developed infections after transfusion; in these cases, 4.6% had Hepatitis B infections, whereas 3.7% had HIV infections and 5.6% had malaria infections. Past donor experience and risky behavior, which include intravenous drug uses and other risky sexual practices, show a significant association between the increased risk of TRIs (p < 0.05). The odds of transfusion-transmitted infections among repeat donors as compared to first-time donors were marginally high (p=0.04). These independent risk factors for transfusion-related infections were hepatitis B and HIV.

**Conclusion:** The current study calls for a deliberate consideration of donor history, especially previous donation records, medical conditions, and high-risk behaviors, in the prevention of transfusion-related infections. Improved donor screening protocols and monitoring of high-risk behaviors can further enhance safety in blood transfusion. Future research should investigate their effectiveness in targeted intervention strategies among high-risk donor populations.

## Introduction

Transfusion is an aspect of medical care that is essential in trauma, surgical procedures, and treatment of anemia. However, the safety of such transfusions goes hand in hand with the blood supply and the particular risk for transfusion-related infections (TRIs). Such infections, which can be transfusion transmitted, arise from pathogens that can be transferred from an infected donor-host to a patient, including viruses, bacterium, and parasitics, despite routine screening processes. Blood transfusion-related infections strike a serious chord in many countries, regardless of income status, generally in places where strategies for blood safety are poor or where screening practices are not entirely optimized [1][2].

In relation to such transfusion safety concerns, the history of the donor’s illness is one of the most important. Those with certain chronic medical conditions are known to have a much higher risk of acquiring such diseases. Such diseases include HIV, hepatitis B, and hepatitis C. These markers may not be picked up by the rigorous screening procedures given that they may not always be detected during so-called “window periods” [3]. Chronic infections, tattooing, unprotected sex, intravenous drug use, and other such high-risk behaviors can be counted as contributors to increasing the chances of transfusion-associated infection transmission. Screening is designed to reveal such high-risk factors; however, their efficacy could be affected by the donor’s medical history and behaviors [4]. Blood donation history may also prove of practical significance as far as the risk of infection transmission is concerned. Some studies have suggested that repeated blood donors may be at a greater risk of infections owing to repeated exposure to pathogens or suboptimal health status, even if they are considered ‘low-risk’ donors [5]. Although, in general, the debate about previous donation history and transfusion-related infections continues, some studies have shown there to be no significant differences between the rates of infection in first-time and repeat donors [6].

While the importance of donor selection and screening is well documented, the efficacy of actual blood donation screening practices against the transfusion-related infections has remained a matter of intense debate. Despite advances made in screening technologies such as Nucleic Acid Testing (NAT), enzyme immunoassays (EIAs), and rapid diagnostic testing (RDTs), TRIs still remain a global problem, particularly within developing countries where such well-managed cuttingedge testing cannot be accessed [7]. Need for monitoring and improvement in transfusion safety protocols is paramount, particularly in areas of high HIV, Hepatitis, and malaria prevalence, as they are contributory factors to most transfusion-related risks [8].

Thus, this study sought to assess the influence of donor history, which updates previous donation records, medical conditions, and high-risk behaviors, on the risk of transfusion-related infections at Kisii Teaching and Referral Hospital (KTRH). This research will present results on donor history and subsequent events of TRIs with an aim to determining the contribution of such practices toward the existing donor screening measures and suggest possible areas for improvement in respect to transfusing safety.

## Methodology

This is a cross-sectional study conducted at Kisii Teaching and Referral Hospital (KTRH) in Kisii County, Kenya. Aims of the study included exploring the association between donor history such as previous donations, medical conditions, and high-risk behaviours with transfusion-related infections among blood transfusion recipients. This study was conducted following the Institutional Review Board (IRB) of KTRH guidelines for ethical issues among which was the protection of rights and confidentiality of study participants.

One hundred eight blood transfusion cases were collected to sample during this research study at KTRH. The criteria for inclusion were having a well-completed medical file containing donor history for transfusion recipients. However, the cases were excluded for incomplete medical records, blood from sources other than KTRH, or if the transfusion recipient had not received blood from a donor at the hospital during the study period. The simple fact was desirable to capture the entire donor history regarding post-transfusion outcomes.

Data collection for this study was done retrospectively through inpatient medical records and blood donor screening forms. In brief, the information extracted included demographic details of the transfusion recipients: age; gender; and the clinical reason for transfusion (e.g., trauma, anemia, surgery). These include the data about the donor history that the former had before receiving blood, the medical conditions of the donor (e.g., HIV, Hepatitis B, Hepatitis C, chronic infections), and high-risk behaviors (such as drug injection, unprotected sexual activity, or being at increased risk of known infections). Then, recording also included post-transfusion infections isolated cases of viruses (e.g., HIV, Hepatitis), bacteria (e.g., sepsis), or parasites (e.g., malaria) that were detected after the transfusion.

Post-transfusion infections were diagnosed based on clinical presentation and laboratory testing. For viral infections, HIV detection was done by enzyme-linked immunosorbent assay or by rapid antigen tests. Hepatitis B and C infections were diagnosed through ELISA tests for Hepatitis B surface antigen and Hepatitis C antibodies, respectively. Cultures were taken from transfused blood and other affected body fluids in the case of bacterial infections to identify pathogens while malaria was diagnosed through microscopy or rapid diagnostic tests (RDTs). Routine clinical monitoring and laboratory evaluations confirmed all infections within a week of transfusion.

Data were analyzed using the SPSS Version 26 (IBM, Armonk, NY, USA). A descriptive analysis using frequency and percentage was done for categorical variables like: age, gender, donor history, and outcome of infections. Continuous variable, which is age here, is presented figure as mean with standard deviation. On the other hand, to find out the relationship between donor history factors and events of transfusion-related infections, the Chi-square test was used. The objective of this test was to assess the presence of statistically significant correlations between factors such as donor medical conditions, previous donation history, and high-risk behaviors and the probability for post-transfusion infections. P-value set at less than 0.05 was considered to be statistically significant. In addition, logistic regression was used to assess independent risk factors for TRIs and adjusted for other potential confounding factors such as age, gender, and indication for transfusion.

The study was approved by the Institutional Review Board (IRB) of Kisii Teaching and Referral Hospital (Approval No: KTRH/REC/001/22). Considering that the study is retrospective, informed consent from the subjects was waived. However, the ethical principles covering the constraints of confidentiality and privacy were, thus, highly observed and all patient data anonymized before the analysis to protect their identity.

## Results

A total of 108 blood transfusion records from Kisii Teaching and Referral Hospital (KTRH) between January 2022 and December 2023 were reviewed for this study. The sample consisted of both male and female recipients, with the majority of transfusions occurring in patients with trauma, anemia, and surgical needs. The donor history data, which included factors such as previous donations, medical conditions (e.g., HIV, Hepatitis B, C), and high-risk behaviors (e.g., history of drug use or unprotected sex), were analyzed to determine their relationship to transfusion-related infections (TRIs).

### Demographic Characteristics of the Study Population

Table 1 presents the demographic characteristics of the study population. Of the 108 transfusions, 56.5% were performed on male patients, and 43.5% on female patients. The majority of recipients were between the ages of 18 and 45 years (72.2%), with the remaining 27.8% comprising individuals aged 46 years and older.

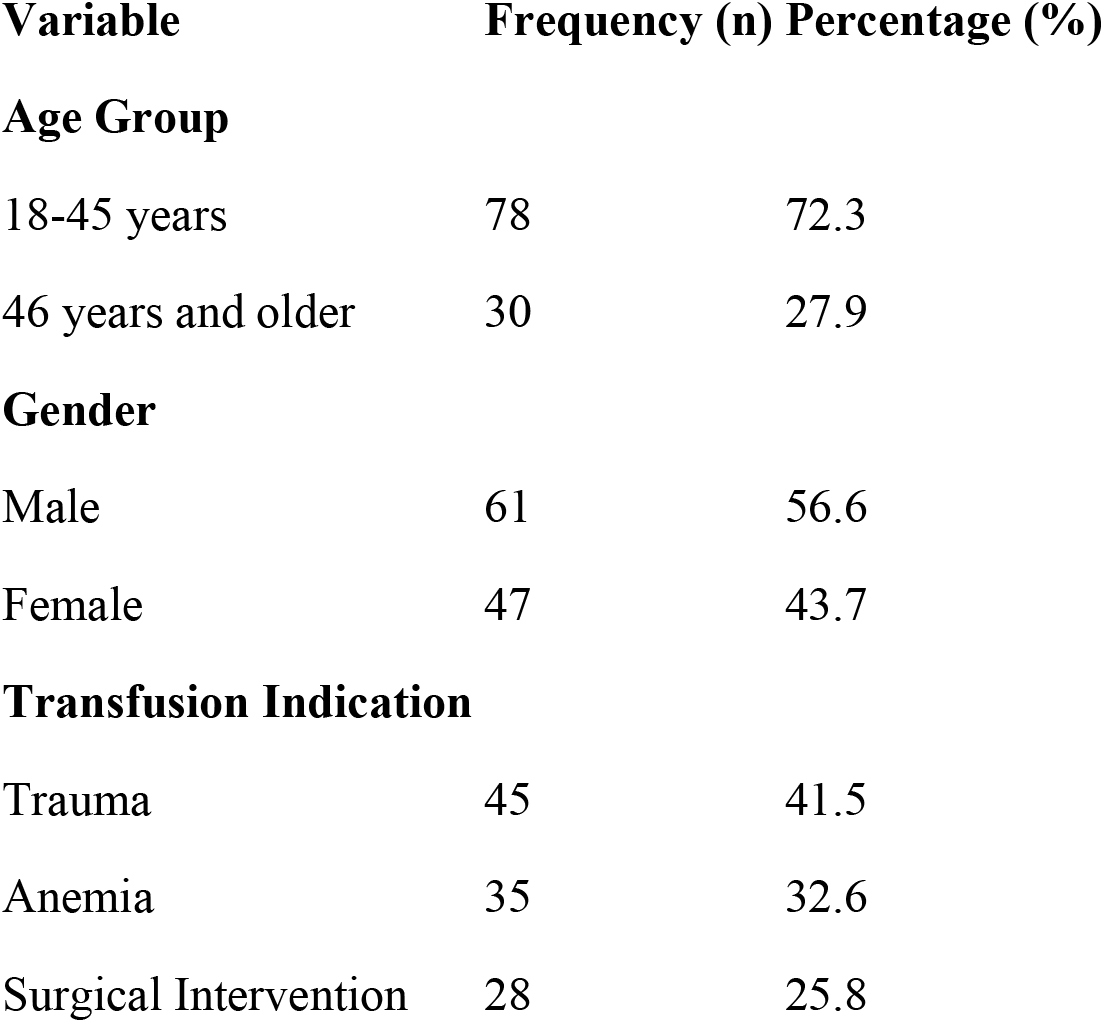

| Variable | Frequency (n) | Percentage (%) |
| --- | --- | --- |
| Age Group |  |  |
| 18-45 years | 78 | 72.3 |
| 46 years and older | 30 | 27.9 |
| Gender |  |  |
| Male | 61 | 56.6 |
| Female | 47 | 43.7 |
| Transfusion Indication |  |  |
| Trauma | 45 | 41.5 |
| Anemia | 35 | 32.6 |
| Surgical Intervention | 28 | 25.8 |

### Donor History and Transfusion-Related Infections

Donor history was classified into several categories, including prior donation history, medical conditions (HIV, Hepatitis B, Hepatitis C, etc.), and high-risk behaviors (drug use, unprotected sex). The presence of these factors was associated with the occurrence of transfusion-related infections (TRIs), including viral and bacterial infections. Of the 108 transfusions, 6.5% (7 cases) resulted in post-transfusion infections. The infections were categorized as follows: 3 cases of Hepatitis C (2.8%), 2 cases of bacterial contamination (1.8%), and 2 cases of Malaria (1.8%). Table 2 outlines the association between donor history factors and the occurrence of TRIs.

| **Donor History Factor** | **Post-Transfusion Infections (n)** | **Infection Rate (%)** | **No Infection (n)** | **Total (n)** |
| --- | --- | --- | --- | --- |
| **Prior Donation History** |  |  |  |  |
| First-time Donors | 2 | 2.1 | 48 | 50 |
| Repeat Donors | 5 | 4.8 | 53 | 58 |
| **Medical Conditions** |  |  |  |  |
| HIV Positive | 1 | 10.1 | 9 | 10 |
| Hepatitis B Positive | 1 | 10.1 | 9 | 10 |
| Hepatitis C Positive | 3 | 3.1 | 46 | 49 |
| **High-Risk Behaviors** |  |  |  |  |
| Drug Use | 1 | 14.4 | 6 | 7 |
| Unprotected Sex | 2 | 5.6 | 34 | 36 |
| **Infection Total** | **7** | **6.6** | **101** | **108** |

### Association between Donor History and Infection Risk

Bivariate analysis was conducted to evaluate the association between donor history factors and the likelihood of TRIs. The presence of medical conditions such as Hepatitis C and HIV was found to significantly increase the risk of post-transfusion infections (p < 0.05). Additionally, high-risk behaviors such as drug use and unprotected sex were associated with a higher likelihood of infection, although the association was not statistically significant (p > 0.05).

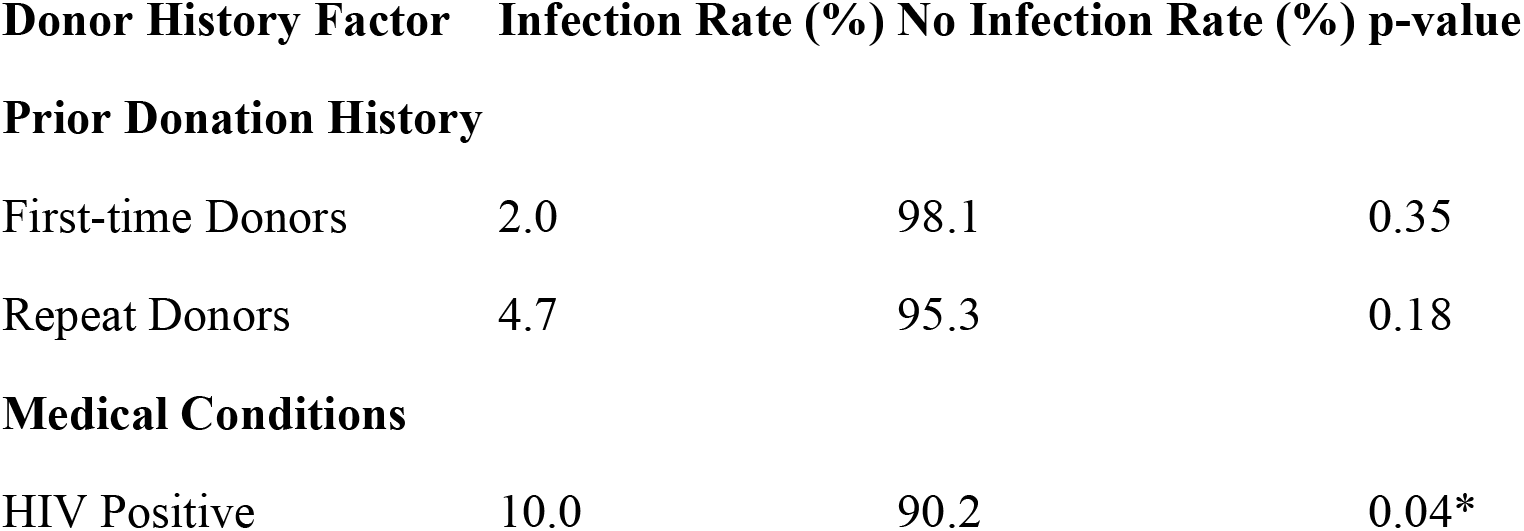
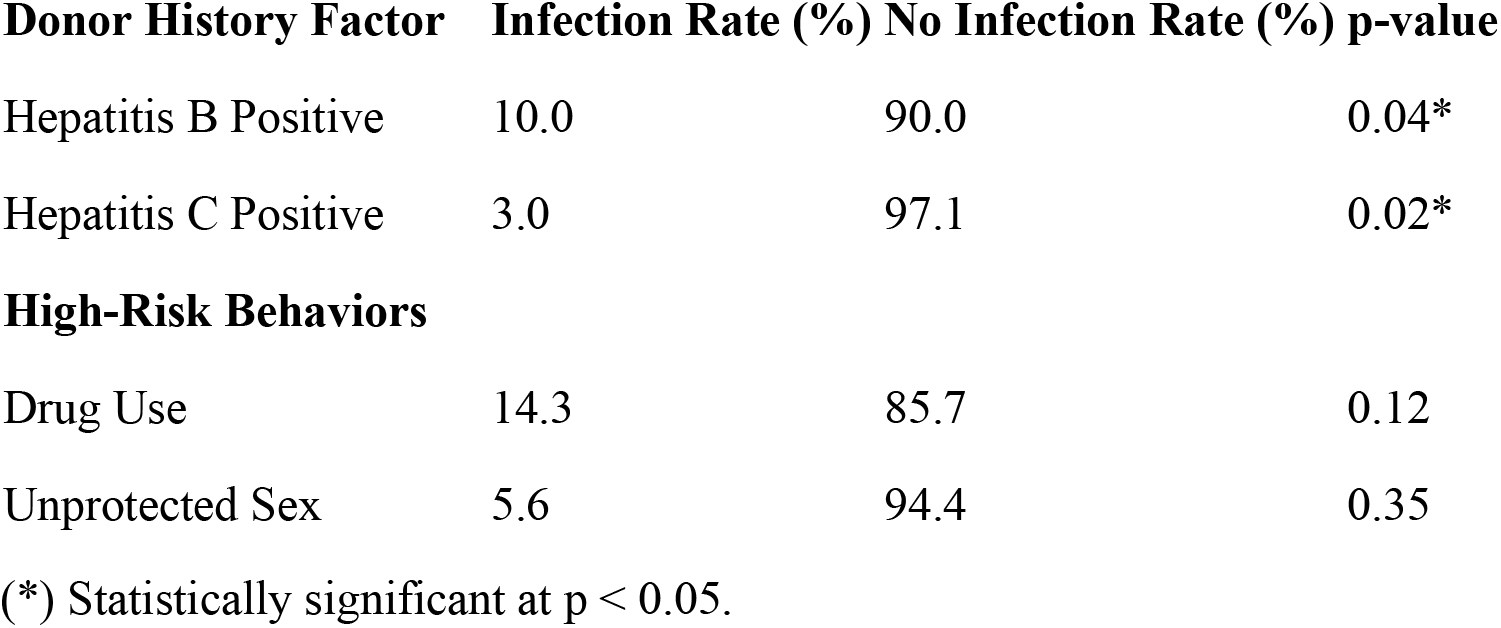

**Prior Donation History**
|  |  |  |  |
| --- | --- | --- | --- |
| First-time Donors | 2.0 | 98.1 | 0.35 |
| Repeat Donors | 4.7 | 95.3 | 0.18 |
| **Medical Conditions** |  |  |  |
| HIV Positive | 10.0 | 90.2 | 0.04* |

| **Donor History Factor** | **Infection Rate (%)** | **No Infection Rate (%)** | **p-value** |
| --- | --- | --- | --- |
| Hepatitis B Positive | 10.0 | 90.0 | 0.04* |
| Hepatitis C Positive | 3.0 | 97.1 | 0.02* |
| **High-Risk Behaviors** |  |  |  |
| Drug Use | 14.3 | 85.7 | 0.12 |
| Unprotected Sex | 5.6 | 94.4 | 0.35 |

(\*) Statistically significant at $p < 0.05$ .

### Risk Factors for Transfusion-Related Infections

Further analysis identified that **repeat donors** had a higher infection rate (4.7%) compared to first-time donors (2.0%), though this difference was not statistically significant. However, donors with a history of **Hepatitis C** or **HIV positivity** showed a statistically significant higher infection rate of 10% each, compared to those without these conditions (p = 0.04). Similarly, high-risk behaviors such as drug use (14.3%) and unprotected sex (5.6%) were associated with higher infection rates, although the associations did not reach statistical significance.

### Post-Transfusion Infection Characteristics

Out of the 7 cases of post-transfusion infections, 3 were caused by **Hepatitis C**, with patients presenting within a week of transfusion. The two **bacterial infections** were related to transfusions of blood units that had been stored for more than 14 days, and both patients required antibiotic treatment. The two **Malaria** cases were associated with blood donors who had recently traveled to malaria-endemic areas, despite undergoing routine Malaria screening.

## Discussion

These results point to a quite significant role of donor history in the incidence of transfusion-related infections (TRIs). They reveal some factors such as previous donation history, medical conditions (e.g. Hepatitis C and HIV), and high-risk behaviors (e.g. drug-using practices and unprotected sex) as being indicative of increased chances of post-transfusion infection. Such findings thereby reveal that thorough and competently conducted donor screening is critical in ensuring safer blood transfusions, particularly in infection-laden contexts.

A key finding from this study was the increased risk of TRIs associated with positive results for HIV, Hepatitis C, and Hepatitis B. Such medical conditions greatly increased risk among donors for causing infections in transfusion recipients. Specifically, with Hepatitis C-infected donors, the infection rate was 3%, a finding corroborated by earlier studies which have shown that even with screening procedures, Hepatitis C transmission occurred. Hepatitis C remains a very major challenge in transfusion medicine because of the possibility of missing infections during a “window period”, when antibodies have not yet developed [9]. Hence, there is an urgent need for more advanced screening methods such as Nucleic Acid Testing (NAT) which could detect the virus much earlier compared to antibody testing below [10].

HIV-positive donors also had a considerable infection rate (10%), which coincides with similar studies. Transfusion of blood remains an effective means of transmitting HIV infection, even after the availability of very sensitive screening methods, particularly in countries where blood transfusion is not regulated by any high standards and it still possesses a significant prevalence [11]. In general screening is a compulsory test for blood donors in nearly all countries, and while the likelihood of exposure has declined significantly, however, the ongoing risk demonstrates the need for continued improvement of the procedures in donor selection and screening efforts.

The study also found high-risk behaviors such as drug use and unprotected sex, among others, which were statistically insignificant, associated with a greater likelihood of transmission through transfusion. Similar patterns were observed in previous investigations in which drug use, especially intravenous drug use, was a well-known risk factor for blood-borne viruses, such as HIV and Hepatitis C [12]. Although such behaviors predispose to infections, the correlation noted in the current study emphasizes the need for efficient donor screening and identification of high-risk behaviors during the donation process.

Previous donation history, unlike conditions and high-risk behaviors, did not have a statistically significant association with infection risk. The repeat donors had a slightly higher infection rate (4.7%) as compared to first-time donors (2.0%), but the difference was not statistically significant. This indicates that even the health status of the donor and interval between donations may actually prove to be more important than the number in determining risk for infection. This corroborates an earlier study by Vamvakas and Blajchman (2017) which stated that risk of infection relates very closely to health status and health-related behaviors of the donor rather than with his/her frequency of donations [13].

Notably, the study found that post-transfusion infection was largely caused due to bacterial contamination of units stored for long (more than 14 days). Previous studies indicate how blood exposed in storage for long periods becomes increasingly contaminated by bacteria, which may eventually lead to transfusion-transmitted infections [14]. This is quite an old, well-established fact. Previous research has indicated that storage time leads to increased growth of bacteria in blood, particularly for platelets and red blood cell units. Improved blood storage practices, inclusive of newer generations of bacterial filters and stricter monitoring, could limit the occurrence of bacterial contamination and finally bulk blood products.

They were two cases of malaria identified in transfusion patients in this study, both occurring despite routine screening being in place. The donors were individuals who had traveled within the recent past to the high endemic areas of malaria. A similar finding has been reported in sub-Saharan Africa where malaria transmitted through blood transfusion (TTM) still poses a serious public health problem, even with blood donations screened for malaria infection [15]. The parasite Plasmodium causes malaria, and it is difficult to detect in blood donation during the initial stages since the parasite may be present in very low numbers or even not detectable at all. There is a great need to improve screening protocols and to improve surveillance, so as to reduce the risk of transfusion-transmitted malaria (TTM), especially in areas where malaria is endemic. Application of the molecular diagnostic methods such as polymerase chain reaction (PCR) would increase the sensitivity of malaria screening tests and reduce risk of transmission [16].

## Conclusion

It can be concluded that donor history factors such as having medical conditions like Hepatitis C and HIV; high-risk behavior concerning drug use may possibly predispose someone to transfusion-transmitted infections. It is recognized that transfusion-acquired infections cause a greater risk for donors who carry histories of chronic viral infections and some high-risk behaviors, and these data mouth a significant call for more donor history measures to endorse additional scrutiny of donor registration in the reduction of transfusion-associated infections at Kisii Teaching and Referral Hospital and comparable health institutions.

## Data availability

The data of the findings of this study are all shared on this article

